# Frequency of Depressive Symptoms and Suicidal Ideation Among University Students Before and After the COVID-19 Pandemic

**DOI:** 10.1101/2024.03.14.24304160

**Authors:** Mélissa Macalli, Laura Castel, Hélène Jacqmin-Gadda, Marie Tournier, Cédric Galéra, Edwige Pereira, Christophe Tzourio

## Abstract

**Background:** The COVID-19 pandemic and lockdown have had negative effects on students’ mental health. However, little information is available regarding the frequencies of depressive symptoms and suicidal ideation during the post-pandemic period. We aimed to compare prevalence rates of depressive symptoms and suicidal ideation among university students, before versus after the COVID-19 pandemic.

**Methods:** In this comparative study, 4464 students were recruited during the pre-COVID-19 pandemic period (2013-2020) and 1768 students, during the post-COVID-19 pandemic period (2022-2023). Standardized frequencies of depressive symptoms and suicidal ideation were compared between the two time periods. Adjusted logistic regression models were used to assess the association between the pandemic period (with the pre-COVID-19 period as reference) and depressive symptoms and suicidal ideation.

**Results:** Compared to participants from the pre-pandemic sample, participants from the post-pandemic sample had higher standardized rates of depressive symptoms (40.6% vs 25.6%) and suicidal ideation (29.3% vs 21.1%). With adjustment for age, gender, university campus, scholarship, and past history of depression and suicide attempt, students in the post-pandemic period experienced more depressive symptoms (aOR, 1.89; 95% CI, 1.67-2.13) and suicidal ideation (aOR, 1.46; 95% CI, 1.28-1.67) compared to students in the pre-COVID-19 era.

**Limitations:** The main limitations were self-selection bias and information bias due to potential over-reporting linked to media coverage, as well as measures of past history of depression and suicide attempt across the lifespan.

**Conclusions:** These findings reveal an alarming deterioration of students’ mental health in the post-pandemic period compared to the pre-pandemic era. Pending replications in others countries, these results suggest that it is crucial to continue monitoring students’ mental health, strengthening communication on this topic, and reinforcing university mental healthcare systems.

## Introduction

Although some studies showed that suicidal risk was stable or diminished during the pandemic (Appleby et al., 2021; Lopez et al.2021), other studies highlighted an increase in mental health problems and suicidality in different populations and age groups (Ettman et al., 2020; Viner et al., 2022; Giannouli et al., 2023). However, there are few recent data in the literature on the post-pandemic period, even for at-risk groups (Wang et al., 2022), e.g., university students. Before the COVID-19 pandemic, university students were known to exhibit a high prevalence of mental health problems, including depressive symptoms and suicidal ideation, in most countries (Auerbach et al., 2018), including France (Macalli et al., 2020). Several factors serve as an explanation as to why university students are such a vulnerable group to mental health disorders: academic stress, separation and individuation from their family, first onset of mental health or substance use problems. This pre-existing vulnerability was exacerbated by the COVID-19 pandemic and the associated lockdowns (Arsandaux et al., 2021; Wathelet et al., 2020). Thus, a previous study showed that students were more likely to have high scores of depressive symptoms and anxiety more frequently than non-students during the lockdown (Macalli et al., 2021). Furthermore, a study conducted among 664 university students in Germany showed that the rate of students suffering from suicidal ideation was twice as high in 2020 than in previous years from 2016 to 2019 (Brailovskaia et al., 2021). However, to our knowledge, few studies have assessed university students’ mental health situation during the current post-pandemic period in comparison to the pre-pandemic period. In the present study, we assessed and compared the prevalence rates of depressive symptoms and suicidal ideation among French university students before versus after the COVID-19 pandemic.

## Methods

### Study Design and Sample

We conducted a comparative analysis using baseline data from two studies of French student volunteers: i-Share and Prisme, which both focused on students’ health and its determinants. These studies used similar recruitment methods (social networks, mailings, meetings at stands and in lecture halls, and a peer-to-peer approach), eligibility criteria (officially registered at a university or higher education institute, at least 18 years of age, and provided informed consent for participation), data collection method (online questionnaires), and measures of mental health parameters. In i-Share, students were enrolled from 2013 to 2020, mainly at Bordeaux university, but also at other French universities. Students enrolled in the i-Share cohort after March 17, 2020 (date of first lockdown in France) were not included in the current study. In Prisme, participants were recruited from September 2022 to February 2023 at Bordeaux university only. To optimize comparability of samples, we restricted the i-Share sample to students from Bordeaux university (pre-COVID-19 pandemic sample, N = 4464). This sample was compared to the Prisme sample (post-COVID-19 pandemic sample, N = 1768). Regulatory approvals were obtained for both studies. The National Commission for Information Technology and Liberties (CNIL) [DR-2013-019] approved the i-Share study. The regional south-east VI ethics committee (*Comite de Protection des Personnes Sud Est VI)* approved the Prisme study (under the National Number 2022-A00666-37). No compensation was paid for participation in either study.

### Measures

Depressive symptoms were measured using the French version of the 9-item Patient Health Questionnaire (PHQ-9), the most commonly used tool for screening for depression (Levis et al., 2019). We used a validated cut-off of 10 to define the presence of depressive symptoms (Kroenke et al., 2001). Suicidal ideation was measured using the following question: “In the past 12 months, have you ever had suicidal thoughts?” In our analyses, we considered the following variables: COVID-19 pandemic period (pre- and post-pandemic), age, gender, university campus (to inform on study fields), scholarship (allocated on social and economic criteria), year of study, perceived stress (measured by the Perceived stress scale-4), and past history of depression and suicide attempt.

### Statistical Analysis

We first compared the pre-pandemic and post-pandemic samples in terms of mental health and socio-demographic characteristics. To obtain more precise estimates of mental health variables, by accounting for the structural differences of the samples, we re-estimated these variables after standardizing the post-pandemic sample structure based on the pre-pandemic sample. To this end, we used the raking ratio method with three margin variables—namely, gender, age, and university campus (macro SAS CALMAR). Finally, to assess the association between the pandemic period (post-pandemic versus pre-pandemic as reference) and the two outcomes, i.e., depressive symptoms and suicidal ideation, we used multivariable logistic regression models for each outcome. Multivariable logistic regression models were performed for the total sample (pre-epidemic and post-epidemic without raking): 1) to estimate the probability of occurrence of depressive symptoms and suicidal ideation among students in the post-epidemic period compared with students in the pre-epidemic period; 2) to conduct a sensitivity analysis adjusted for confounding factors (scholarship, past depression (for depressive symptoms), or suicide attempt (for suicidal ideation)). In secondary analyses, to clarify the comparison between the post-COVID period (from September 2022 to February 2023) with the preceding seven years’ pre-era, we repeated the analyses limiting the pre-pandemic sample to the three previous years before the pandemic (2018-2020).

## Results

Participants in the pre-pandemic sample were slightly older (mean age, 20.4 years; SD = 2.6; N = 4464) than participants in the post-pandemic sample (mean age, 19.9 years; SD = 2.7; N = 1768) (Table 1). The two samples had similar gender distribution (77.9% women in the pre-pandemic, and 80.8% in the post-pandemic sample), and socio-economic status measured by scholarship (41.8% in the pre-pandemic, and 45.1% in the post-pandemic sample). Compared to participants in the pre-pandemic sample, participants in the post-pandemic sample had higher rates of depressive symptoms (43.3% vs. 25.6%) and suicidal ideation (30.6% vs 21.1%). Other mental health parameters (high perceived stress, and past history of depression and suicide attempt) were also higher in the post-pandemic sample than in the pre-pandemic sample (Table 1).

**Table 1.** Comparison of Key Variables Between Pre- and Post-COVID-19 Samples.

| Characteristic, N (%) | Pre-pandemic sample (N = 4664) | Post-pandemic sample (N = 1768) | Standardized post-pandemic sample (N = 4664) |
| --- | --- | --- | --- |
| **Female** | 3634 (77.9) | 1429 (80.8) | 3634 (77.9) |
| **Age, mean (SD)** | 20.4 (2.6) | 19.9 (2.7) | 20.4 (2.9) |
| **University campus** |  |  |  |
| Bordeaux | 3651 (78.3) | 922 (52.1) | 3651 (78.3) |
| Bordeaux-Montaigne | 769 (16.5) | 525 (29.7) | 769 (16.5) |
| Other or unknown | 244 (5.2) | 321 (18.2) | 244 (5.2) |
| **First year of study <sup>a</sup>** | 1785 (38.6) | 694 (39.3) | 1525 (32.7) |
| **Scholarship <sup>b</sup>** | 1949 (41.8) | 798 (45.1) | 2102 (45.1) |
| **High perceived stress <sup>c</sup>** | 1893 (40.6) | 1033 (58.5) | 2556 (54.9) |
| **Past history of depression <sup>d</sup>** | 489 (10.5) | 327 (18.5) | 873 (18.7) |

| Characteristic, N (%) | Pre-pandemic sample<br>(N = 4664) | Post-pandemic sample<br>(N = 1768) | Standardized post-pandemic sample (N = 4664) |
| --- | --- | --- | --- |
| **Past history of suicide attempt <sup>e</sup>** | 254 (5.4) | 206 (11.7) | 514 (11.0) |
| **Depressive symptoms <sup>f</sup>** | 1194 (25.6) | 765 (43.3) | 1891 (40.6) |
| **Suicidal ideation** | 982 (21.1) | 541 (30.6) | 1366 (29.3) |

<sup>a</sup>Missing data for 38 participants in the pre-epidemic sample.
<sup>b</sup>Missing data for 4 participants in the pre-epidemic sample.
<sup>c</sup>Missing data for 2 participants in the post-epidemic sample, and 5 in the standardized post-epidemic; Score $\geq 8$ used to define high perceived stress, using the PSS-4 scale.
<sup>d</sup>Missing data for 1 participant in the post-epidemic sample, and 3 in the standardized post-epidemic sample.
<sup>e</sup>Missing data for 76 participants in the pre-epidemic sample.
<sup>f</sup>Score $\geq 10$ used to define the presence of depressive symptoms, using the PHQ-9 scale.

After standardizing the post-pandemic sample based on the pre-pandemic sample’s distributions of gender, age, and university campus, we found that the estimated prevalence rates of depressive symptoms (40.6%) and suicidal ideation (29.3%) in the post-pandemic sample were lower than before standardization, but these estimates remained higher than those in the pre-pandemic sample (Table1).

A comparison of socio-demographic characteristics according mental health outcomes in each sample showed that students who reported depressive symptoms and suicidal ideation were more likely female, first year and scholarship students, whatever the period. They were also more likely to present high perceived stress and past history of depression or suicide attempt than those who declared no depressive symptoms nor suicidal ideation (Supplementary material table 1).

We also performed multivariate analyses, with adjustments for age, gender, university campus, scholarship, and past history of depression or suicide attempt. These results showed that compared to participants from the pre-pandemic sample, participants from the post-pandemic sample had an approximately 90% increased risk of depressive symptoms (aOR, 1.89; 95% CI, 1.67-2.13) and an approximately 50% increased risk of suicidal ideation (aOR, 1.46; 95% CI, 1.28-1.67) (Table 2). Female gender (aOR, 1.72; 95% CI, 1.49-2.00), university campus of Bordeaux Montaigne i.e. humanities and social sciences (in reference to Bordeaux campus, i.e. sciences and health section; aOR, 1.39; 95% CI, 1.21-1.59), scholarship (aOR, 1.20; 95% CI, 1.07-1.37), and past history of depression (aOR, 2.95; 95% CI, 2.52-3.46) were associated to a higher probability to report depressive symptoms. Students who reported suicidal ideation were more likely to declare past history of suicide attempt (aOR, 5.22; 95% CI, 4.27-6.39) (Table 2).

**Table 2.**
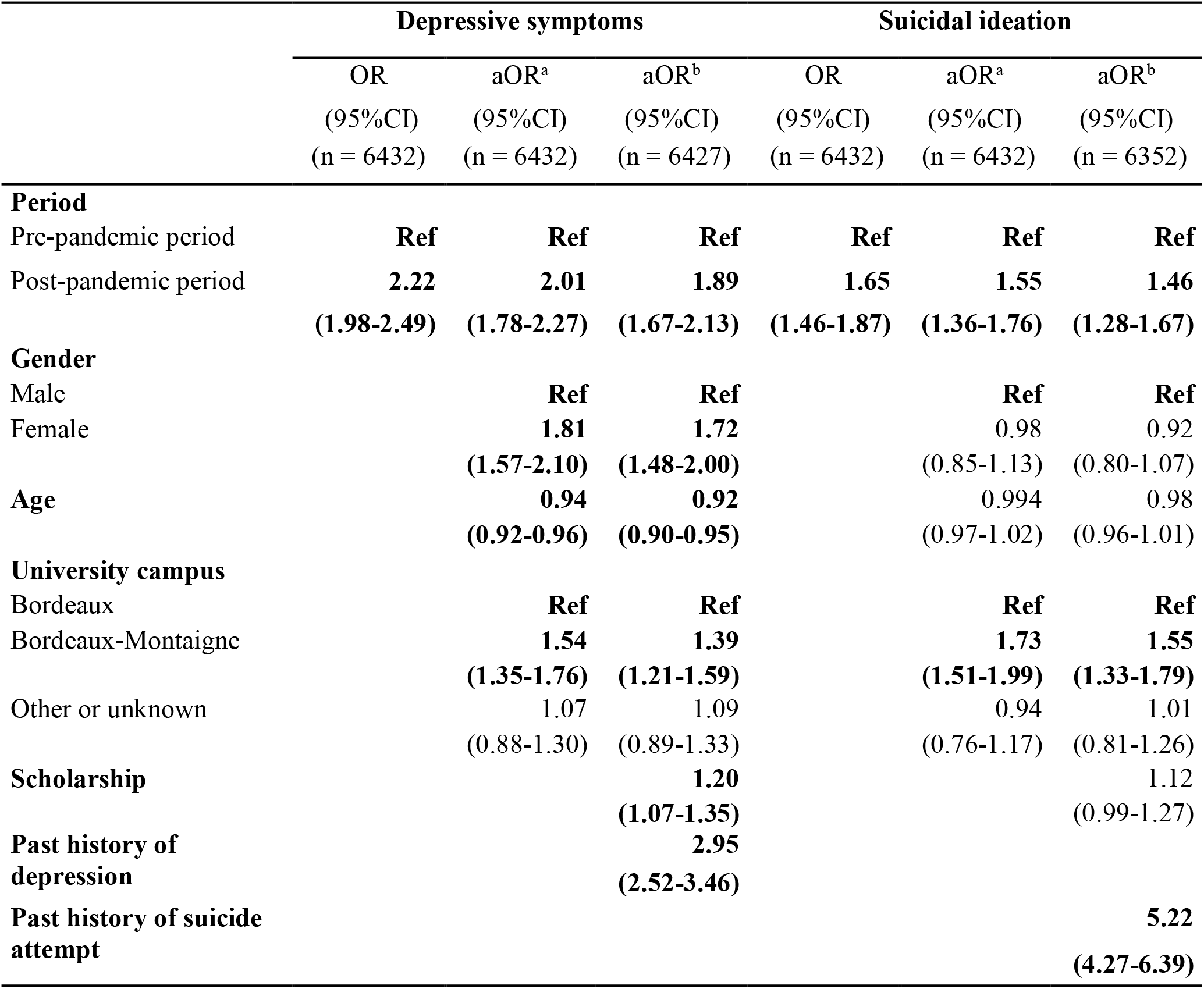
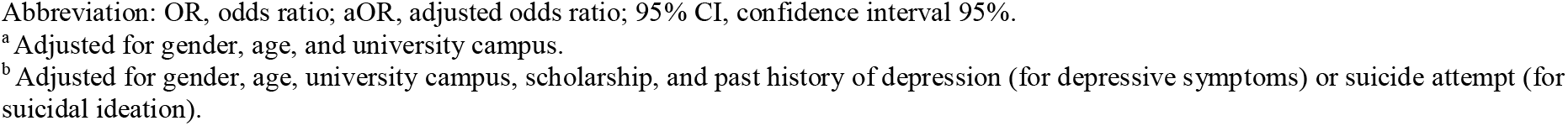
Association Between the Pandemic Period and Depressive Symptoms and Suicidal Ideation.

|  | Depressive symptoms |  |  | Suicidal ideation |  |  |
| --- | --- | --- | --- | --- | --- | --- |
|  | OR<br>(95%CI)<br>(n = 6432) | aOR<sup>a</sup><br>(95%CI)<br>(n = 6432) | aOR<sup>b</sup><br>(95%CI)<br>(n = 6427) | OR<br>(95%CI)<br>(n = 6432) | aOR<sup>a</sup><br>(95%CI)<br>(n = 6432) | aOR<sup>b</sup><br>(95%CI)<br>(n = 6352) |
| **Period** |  |  |  |  |  |  |
| Pre-pandemic period | **Ref** | **Ref** | **Ref** | **Ref** | **Ref** | **Ref** |
| Post-pandemic period | **2.22**<br>(1.98-2.49) | **2.01**<br>(1.78-2.27) | **1.89**<br>(1.67-2.13) | **1.65**<br>(1.46-1.87) | **1.55**<br>(1.36-1.76) | **1.46**<br>(1.28-1.67) |
| **Gender** |  |  |  |  |  |  |
| Male |  | **Ref** | **Ref** |  | **Ref** | **Ref** |
| Female |  | **1.81**<br>(1.57-2.10) | **1.72**<br>(1.48-2.00) |  | **0.98**<br>(0.85-1.13) | **0.92**<br>(0.80-1.07) |
| **Age** |  | **0.94**<br>(0.92-0.96) | **0.92**<br>(0.90-0.95) |  | **0.994**<br>(0.97-1.02) | **0.98**<br>(0.96-1.01) |
| **University campus** |  |  |  |  |  |  |
| Bordeaux |  | **Ref** | **Ref** |  | **Ref** | **Ref** |
| Bordeaux-Montaigne |  | **1.54**<br>(1.35-1.76) | **1.39**<br>(1.21-1.59) |  | **1.73**<br>(1.51-1.99) | **1.55**<br>(1.33-1.79) |
| Other or unknown |  | 1.07<br>(0.88-1.30) | 1.09<br>(0.89-1.33) |  | 0.94<br>(0.76-1.17) | 1.01<br>(0.81-1.26) |
| **Scholarship** |  |  | **1.20**<br>(1.07-1.35) |  |  | **1.12**<br>(0.99-1.27) |
| **Past history of depression** |  |  | **2.95**<br>(2.52-3.46) |  |  |  |
| **Past history of suicide attempt** |  |  |  |  |  | **5.22**<br>(4.27-6.39) |

Abbreviation: OR, odds ratio; aOR, adjusted odds ratio; 95% CI, confidence interval 95%.
<sup>a</sup> Adjusted for gender, age, and university campus.
<sup>b</sup> Adjusted for gender, age, university campus, scholarship, and past history of depression (for depressive symptoms) or suicide attempt (for
suicidal ideation).

The description of prevalences per year in the i-Share cohort from 2013 to 2020 show a slight uptick in depression symptoms and a stability in suicidal ideation, but not a clear trend (Supplementary Material Table 2). In secondary analyses on a subsample (n=569) limited to the three years before the pandemic (2018-2020), participants in the post-pandemic period had higher rates of depressive symptoms (43.3% vs. 28.5%) and suicidal ideation (30.6% vs 21.8%) compared to participants in the pre-era (Supplementary Material Table 3). In multivariate analyses, participants from the post-pandemic sample were more likely to report depressive symptoms (aOR, 1.61; 95% CI, 1.29-2.00) and suicidal ideation (aOR, 1.42; 95% CI, 1.11-1.80) compared to participants from the pre-pandemic sample (Supplementary Material Table 4).

## Discussion

The present results showed that compared to participants from the pre-pandemic sample, those in the post-pandemic sample had higher rates of depressive symptoms and suicidal ideation. Previous studies have shown that young adults, including students, were strongly exposed to depressive disorders during the COVID-19 pandemic (Bliddal et al., 2023; COVID-19 Mental Disorders Collaborators., 2021; Léon et al., 2023). For instance, a study conducted among 884 college students in Northern Ireland and Republic of Ireland from 2019 to 2020 found a large increase in depression by over 10% (McLafferty, et al., 2021). Our findings suggest that this increased frequency of depressive disorders is persisting after the pandemic, rather than returning to normal. A similar observation was reported after the SARS pandemic in 2002, with some studies reporting long-term psychiatric implications, particularly among the survivors (Lee et al., 2007).

As student mental health has steadily declined over the past decade, we can assume that the higher prevalence of depressive symptoms and suicidal ideation in 2022-2023 may be part of the ongoing trend. However, the description of prevalences per year in the i-Share cohort from 2013 to 2020 did not show a regular increase across years. Furthermore, the analyses limited on a subsample of the last three years before the pandemic confirmed our global results suggesting an increase in mental health problems in the post-COVID period rather than a continuum of an existing trajectory.

The literature includes only sparse and inconsistent findings of whether the aggravation of mental health problems during the COVID-19 pandemic also manifested as changes in suicidality. Some studies have shown no change in suicidal ideation at the beginning of the pandemic (Knudsen et al., 2021; Pirkis et al., 2021) or after the lockdown (Danielson et al., 2023), while others have found an increase of suicidal ideation (Huang et al., 2022). In a study conducted among 4693 Canadian university students, Jones et al. observed an initial drop of suicidal ideation followed by an increasing trend, suggesting the possibility of delayed impact (Jones et al., 2023). Likewise, a large study conducted among 44 898 university students found that the prevalence of suicidal ideation increased between the first lockdown (10.6%) in France and August 2021, 15 months later (13.8%) (Wathelet et al., 2022). In this study, in line with our findings, female, students with financial difficulties or psychiatric history, were at risk of mental health problems.

There has been no previous report of the frequency of suicidal ideation among students during the current post-pandemic period in comparison with the pre-era, and our results suggest that there is a possible long-term impact. The aggravation of students’ mental health problems during the post-pandemic period may have various causes, including the economic crisis and persistent anxiety about global warming in this population. Nevertheless, we think that an exacerbation of this magnitude is partly attributable to the COVID-19 pandemic and the lockdowns, which have been traumatic in this population, as they dramatically limited interactions with teachers and peers for several months (Macalli et al., 2021). Some studies revealed a high prevalence of mental health disorders, including anxiety, and depression among students, in the post-Covid era (Chen et al., 2023; Cheng et al., 2023). Contrary to our study, one recent research conducted among Italian university students showed that the freshmen recruited after the pandemic presented less psychological distress than freshmen recruited before the pandemic. However, 78% of the freshmen stated that the pandemic had an impact on their social relationships (Buizza et al., 2023). The WHO revealed that, faced with extended university closures, young people have been left vulnerable to social isolation which can lead to anxiety, uncertainty, loneliness and behavioral problems (WHO, 2022). We can assume that the reconstruction and recovery of certain social and personal skills may take time and lead to some mental exhaustion. In addition, mental health services, already inadequate before the epidemic, may have been overburdened, resulting in ineffective or delayed treatment.

### Strengths and Limitations

Main strengths of this study included the relatively large samples of students (pre-pandemic N = 4664; post-pandemic N = 1768) recruited from the same university, using similar recruitment methods, and using the same validated tools to evaluate mental health. Notably, both studies would have been similarly affected by self-selection bias, which is inherent to community-based studies. Nevertheless, this bias limits the generalizability of our findings. Another potential limitation is the possibility that media coverage of the impact of the pandemic on mental health may have led to over-reporting of mental health disorders during the post-pandemic period. However, although this could have been the case during the pandemic and lockouts periods, it seems rather unlikely that it would have a major impact in the current post-pandemic period, during which these topics have vanished from the media. Because past depression and past suicide attempt were queried across the lifespan in both studies, we cannot use this questionnaire to distinguish whether they occurred in the pre-epidemic period or during the epidemic in the post-Covid sample. Similarly, we cannot determine whether or not past mental disorders are related to current mental health status. However, we did account for this important confounder in our models.

### Conclusions

The present findings reveal an alarming deterioration of students’ mental health in the post-pandemic period compared to in the pre-pandemic period. These results remain to be replicated in other settings and other countries, but they suggest that it is crucial to continue monitoring students’ mental health, strengthening communication on this topic, and reinforcing university mental healthcare systems.

### Data Sharing Statement

In accordance with the General Data Protection Regulation (GDPR), the data sets used and/or analyzed during the current study are available from the corresponding author on reasonable request.

### Authorship contribution statement

MM, EP, HJG, and CT designed the study. LC conducted the statistical analysis. MM and CT wrote the first draft of the manuscript. All authors contributed to editing the manuscript and commenting on the final version.

## Data Availability

All data produced in the present study are available upon reasonable request to the authors

https://research.i-share.fr

## Acknowledgements

The authors are indebted to the Healthy team (Bordeaux Population Health, Inserm) for their contributions, and Kappa Santé/Kap Code teams for their technical expertise. The authors thank all the students that participated in the i-Share and Prisme studies.

